## Supplementary materials for "Susceptibility and infectiousness of SARS-CoV-2 in children versus adults, by variant (wild-type, Alpha, Delta): a systematic review and meta-analysis of household contact studies"

**Online Only Figures**

**eFigure 1: Forest plot for susceptibility showing secondary attack rates among children, adolescents and adults for wild-type.**

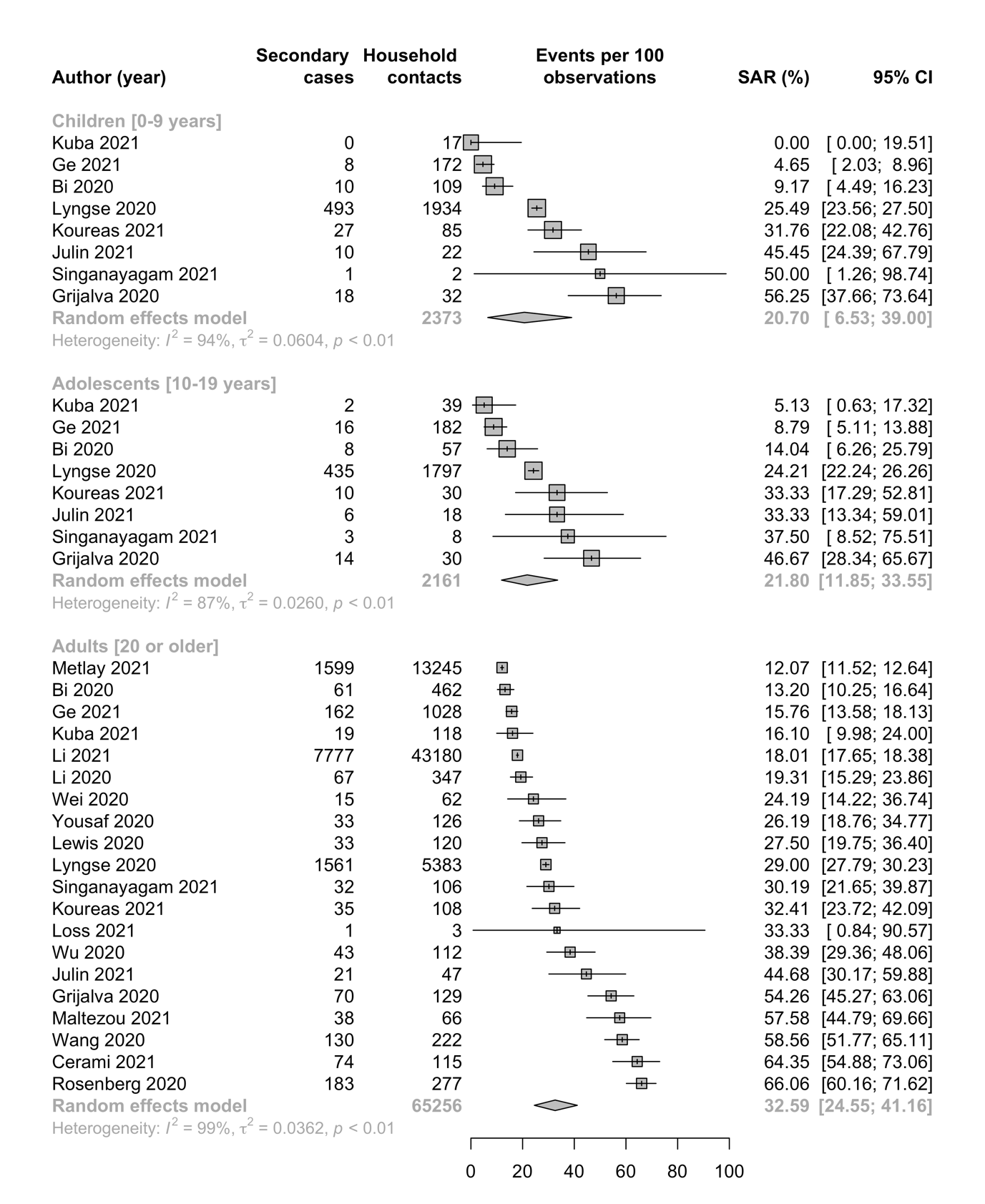

**eFigure 2: Forest plot for susceptibility showing secondary attack rates among children, adolescents, and adults for Alpha**

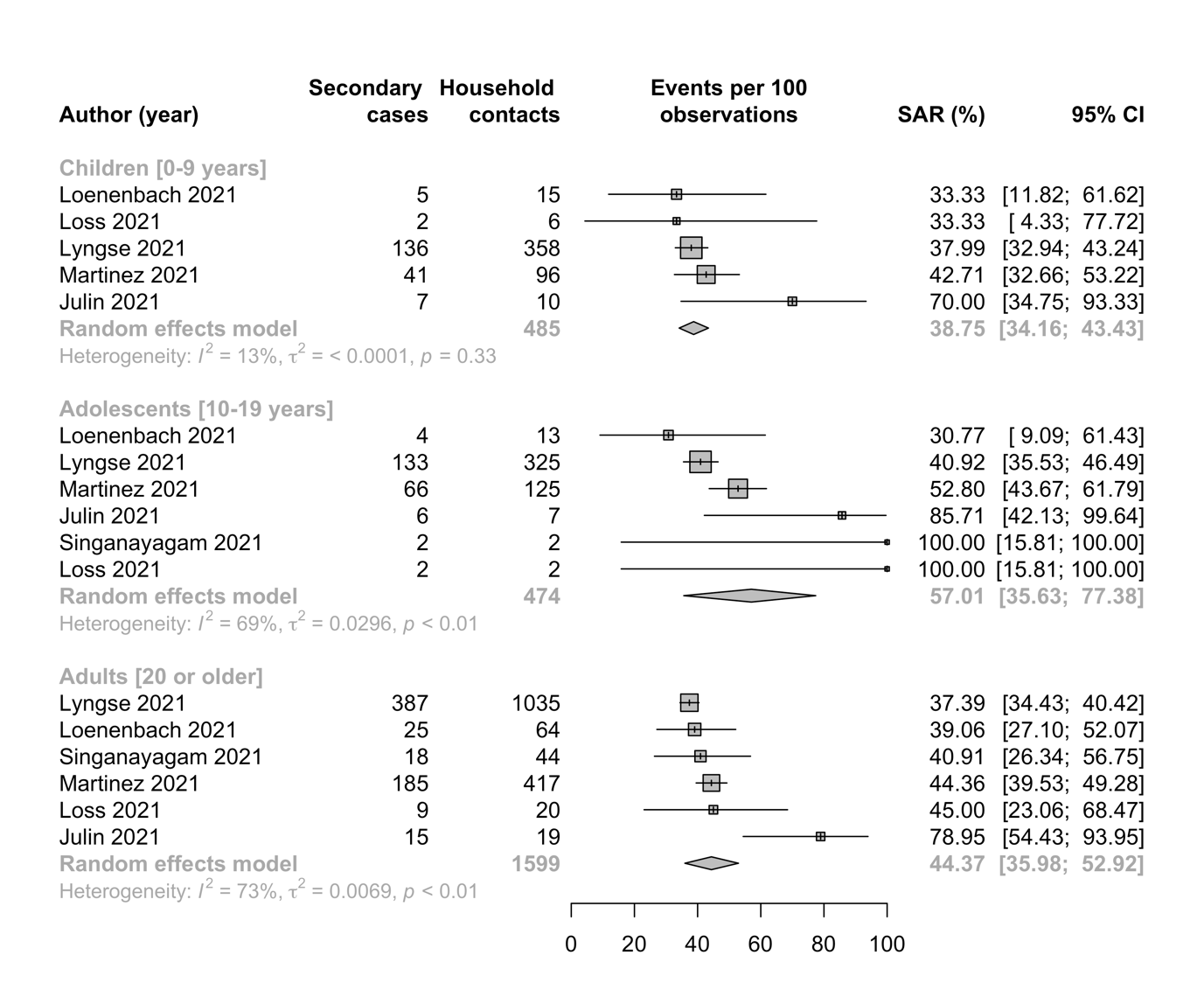

**eFigure 3: Forest plot for susceptibility showing secondary attack rates among children, adolescents, and adults for Delta**

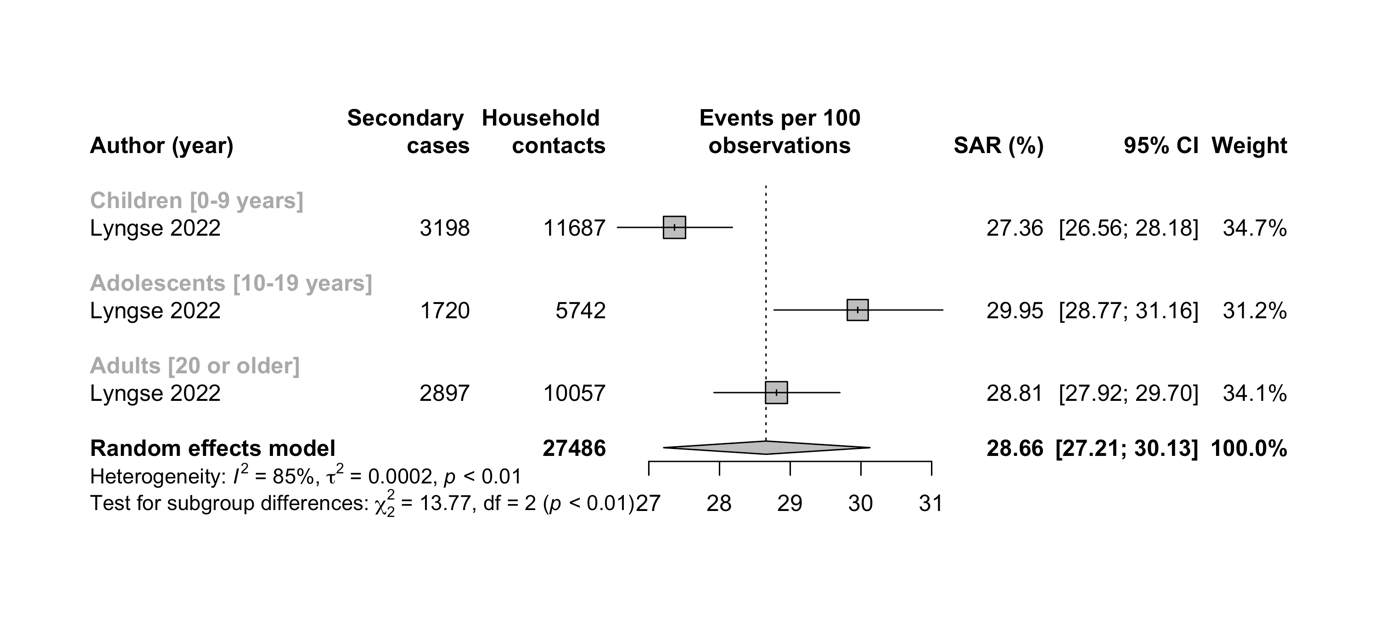

**eFigure 4: Forest plot for infectiousness showing secondary attack rates when children, adolescents and adults were index cases and infected with wild-type SARS-CoV-2.**

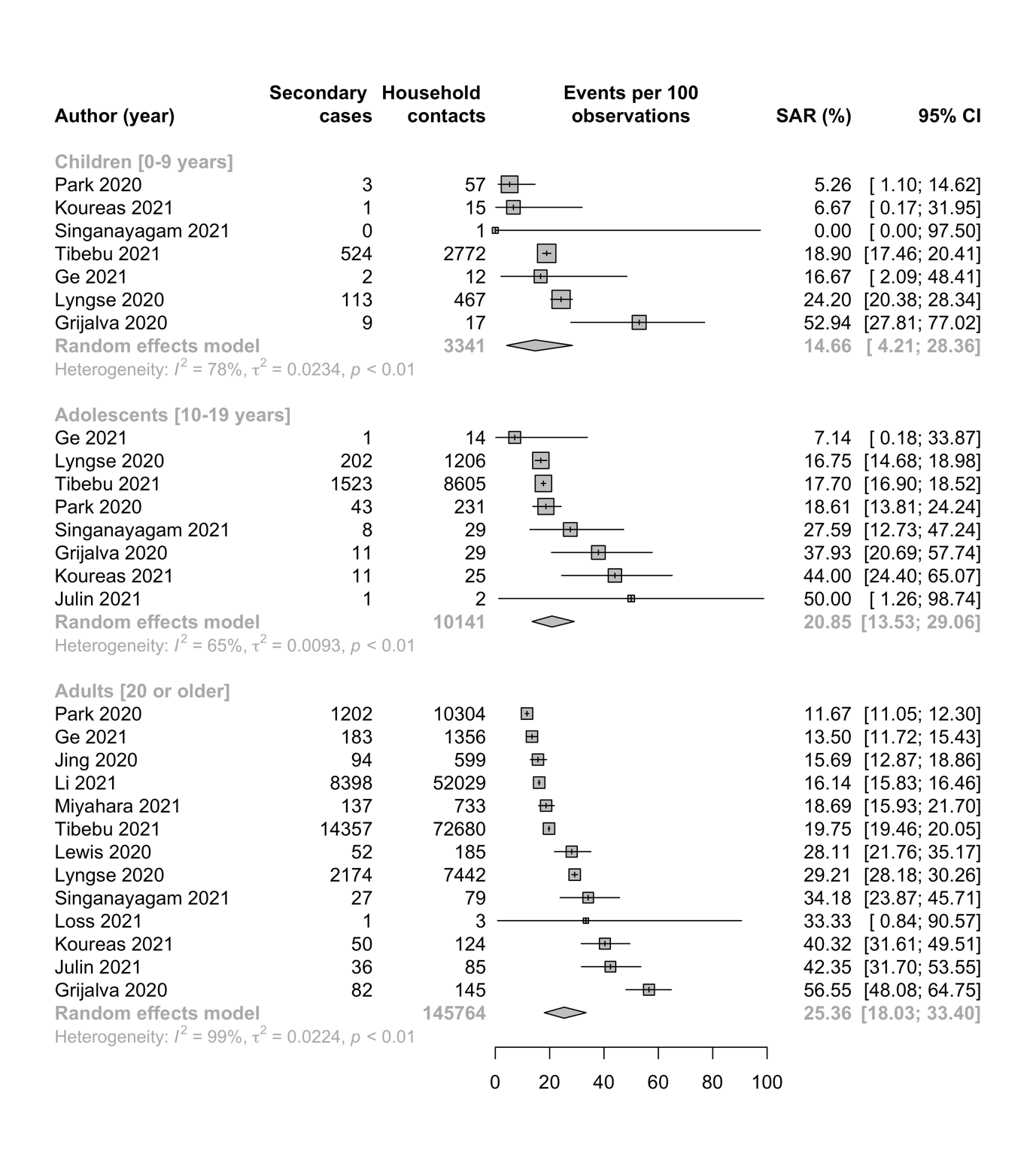

**eFigure 5: Forest plot for infectiousness showing secondary attack rates when children, adolescents and adults were index cases and infected with Alpha variant of SARS-CoV-2.**

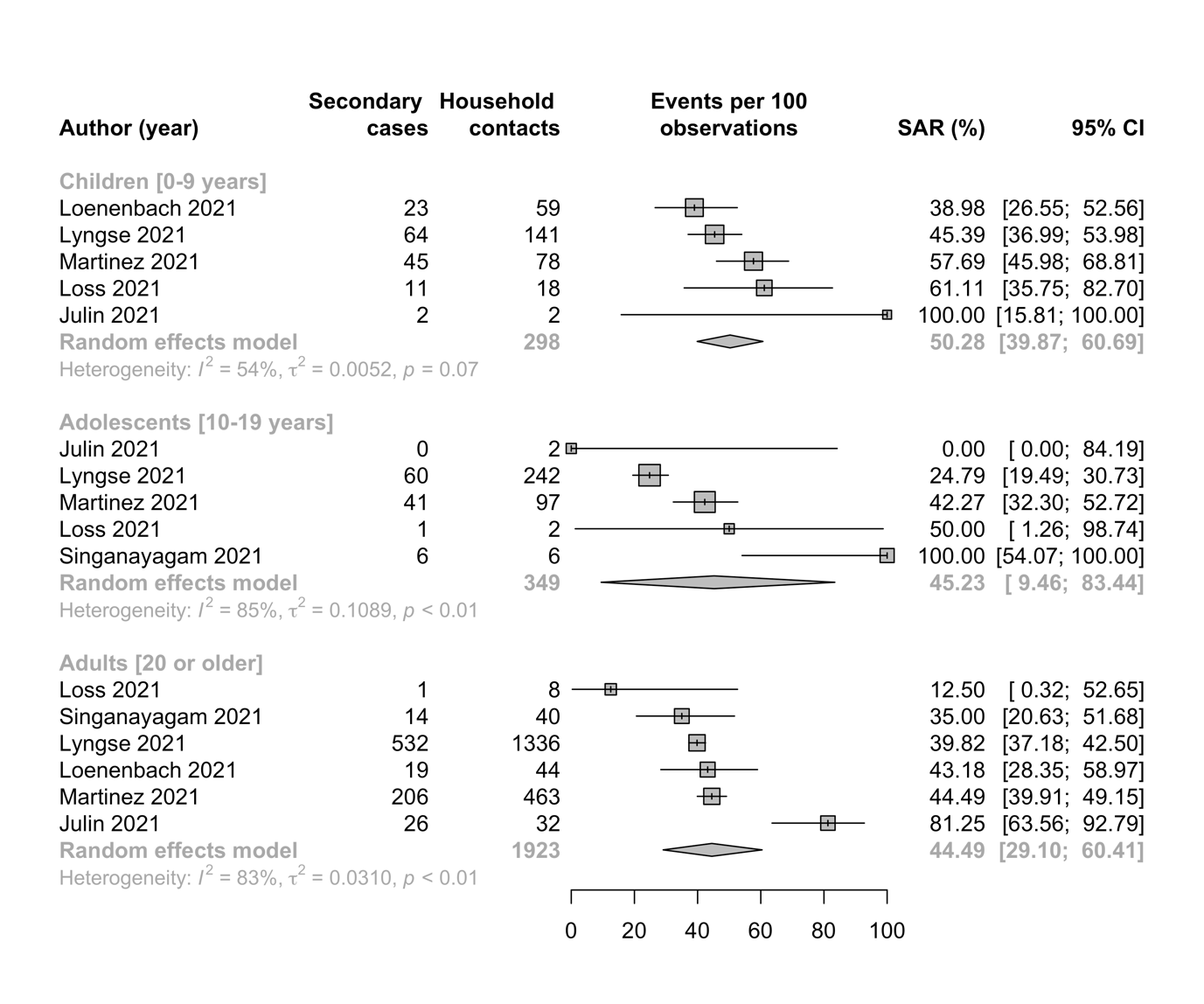

**eFigure 6: Forest plot for infectiousness showing secondary attack rates among children, adolescents, and adults for Delta**

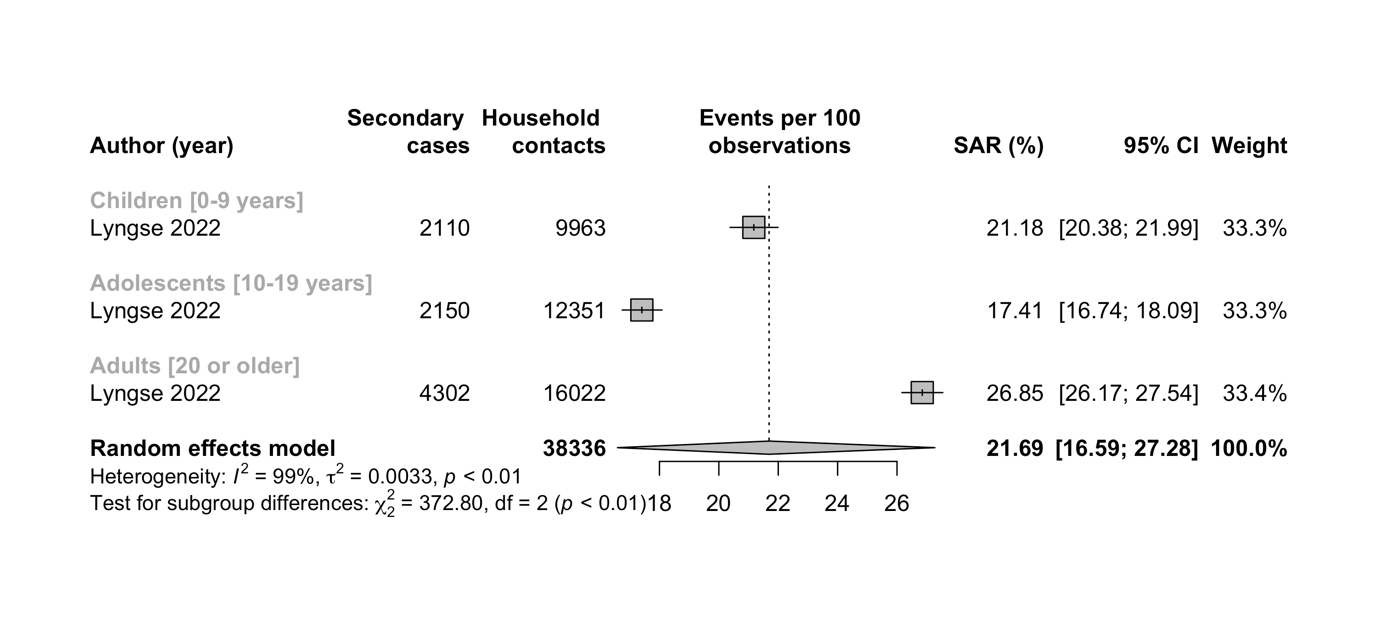

**eTable 1: Summary characteristics of included studies**

| **Study** | **Country of study** | **Study design:** | **Variant type** | **Study period:** | **No of index cases** | **No of contact identified** | **Household SAR (%)** | **Mean age (yrs)** | **% Female** | **Number of tests per contact** | **Follow up duration (days)** | **Case identification** | Case definition index case | Case definition secondary case | Vaccination |
| --- | --- | --- | --- | --- | --- | --- | --- | --- | --- | --- | --- | --- | --- | --- | --- |
| Bi 2020^22^ | China | Retrospective cohort | Wild-type | January 14 to February 12, 2020 | 397 | 1,286 | 11.2 | 45 | 48 | Multiple | 12 plus | Real-time reverse transcription polymerase chain reaction (RT-PCR) of nasal swabs | Community cases | Close contacts were identified through contact tracing of a confirmed case and were defined as those who lived in the same apartment, shared a meal, travelled, or socially interacted with an index case 2 days before symptom onset | Likely all households unvaccinated |
| Cerami 2021^31^ | US | Prospective cohort | Wild-type | April 2020 to November 2020. | 106 | 213 | 0.6 | Not provided | 52 | Multiple | 28 | RT-PCR of nasal swabs | Cases recruited after testing at the Respiratory Diagnostic Center | Cases were detected within 14 days of symptom onset in the index case | Likely all households unvaccinated |
| Ge 2021^32^ | China | Prospective cohort | Wild-type | January 8 to July 30, 2020 | 730 | 8,852 | 10.1 | 46 | 48.8 | Multiple | 14 | RT-PCR using respiratory specimens | Community cases | Active symptomatic follow-up | Likely all households unvaccinated |
| Grijalva 2020^16^ | US | Surveillance data | Wild-type | April–September 2020 | 101 | 191 | 53 | 28 | 54 | Multiple | 14 | RT-PCR of nasal swabs or saliva specimens | Community cases | Active symptomatic follow-up | Likely all households unvaccinated |
| Jing 2020^23^ | China | Retrospective cohort | Wild-type | Jan 7, 2020, and Feb 18, 2020 | 349 | 195 | 12.4 | Not provided | 50 | Multiple | 14 | RT-PCR of nasal swabs | Community cases | Passive identification of secondary cases independent from symptoms | Likely all households unvaccinated |
| Julin 2021^33^ | Norway | Prospective cohort | Wild-type, Alpha | May 2020 to May 2021 | 65 | 200 | 49.6 | 31 | 52 | Multiple | 42 | RT-PCR of Oropharyngeal (OP) samples and neat saliva samples | A >=12 yo case in a household with the first positive test result (“primary case”) and where no other household contact person tested positive on the same day. | Households with vaccinated individuals not eligible. All hhc >=2 years were tested several times until day 14 after test date of primary case irrespective of symptoms. | Households with vaccinated individuals excluded. |
| Koureas 2021^17^ | Greece | Surveillance data | Wild-type | 8 April to 4 June 2020 | 142 | 135 | 38.62 | Not provided | Not provided | Multiple | 14 | RT-PCR of oropharyngeal samples | Community cases | Passive identification of secondary cases independent from symptoms | Likely all households unvaccinated |
| Kuba 2021^34^ | Japan | Surveillance data | Wild-type | February 14 to May 31, 2020 | 78 | 174 | 12.1 | 51.5 | 32 | 1 | 14 | RT-PCR of throat and nasopharyngeal swaps | Community cases | Passive identification of secondary cases independent from symptoms | Likely all households unvaccinated |
| Lewis 2020^35^ | US | Surveillance data | Wild-type | 4 months | 575,071 | 62 | 29 | Not provided | Not provided | Multiple | 14 | RT-PCR of nasopharyngeal or anterior nasal swabs | Community cases | Passive identification of secondary cases independent from symptoms | Likely all households unvaccinated |
| Li 2020^24^ | China | Retrospective cohort | Wild-type | January 1 to February 20, 2020 | 105 | 392 | 18 | 47 | 43.9 | Multiple | 14 | RT-PCR of nasopharyngeal swaps | Community cases | Passive identification of secondary cases independent from symptoms | Likely all households unvaccinated |
| Li 2021^36^ | China | Retrospective cohort | Wild-type | Dec 2, 2019 to April 18, 2020 | 27,101 | 27,101 | 15·6 | 56 | 48 | Not provided | No clearly stated | RT-PCR of respiratory specimens | Community cases | Passive identification of secondary cases independent from symptoms | Likely all households unvaccinated |
| Loenenbach 2021^37^ | Germany | Retrospective cohort | Alpha | 2021 | Not provided | 92 | 37 | Not provided | Not provided | 1 | 14 | PCR test | Cases of an outbreak in a childcare centre | Passive identification of secondary cases independent from symptoms. | Likely all households unvaccinated |
| Loss, 2021^38^ | Germany | Cohort | Alpha, Wild-type | October 2020 to June 2021 | 33 | 45 | 53.3 | Not provided | 61 | Multiple | 12 | RT-PCR | Cases recruited in a study in childcare centres | Active follow-up and systematic, serial testing of household contacts irrespective of symptoms | Adults were asked about their vaccination status in a telephone interview. None of the adults were fully vaccinated (5 individuals in the household cohort had received 1 of 2 shots shortly before the home visits). |
| Lyngse 2020^25^ | Denmark | Register data | Wild-type | February - July 24, 2020 | 6,782 | 6,609 | 17 | Not provided | Not provided | Multiple | 14 | RT-PCR | Primary case was first positive test for SARS-CoV-2. | Secondary cases were defined as those who had a positive test within 14 days of the primary case being tested positive. | Likely all households unvaccinated |
| Lyngse 2021^39^ | Denmark | Register data | Alpha | January 11 to February 7, 2021 | 8,093 | 16,612 | 25 |  | 45 | Multiple | 14 | RT-PCR | Primary case was first positive test for SARS-CoV-2. | Secondary cases were defined as those who had a positive test within 14 days of the primary case being tested positive. | Likely all households unvaccinated |
| Lyngse 2022^46^ | Denmark | Register data | Delta | June 21 to October 25, 2021 | 24,693 | 16,612 | 25 |  | 45 | Multiple | 14 | RT-PCR | The case in a household with the first positive test result (“primary case”) and where no other household contact person tested positive on the same day. | Household contact persons testing positive 1-14 days after test date of the primary case. No information provided if testing based on symptoms or recommended on all household contact persons. More than 70% of all hhc were tested. | Documented and analysed separately for vaccinated/unvaccinated index/primary and hhc. |
| Maltezoua 2021^40^ | Greece | Surveillance data | Wild-type | Febrary 26-May 3, 2020 | 109 household | 109 | 8.8 | Not provided | Not provided | Not stated | 14 | RT-PCR on respiratory specimens | Community cases | Passive identification of secondary household cases independent from symptoms. | Likely all household unvaccinated |
| Martinez 2021^41^ | US | Cohort | Alpha | June 11, 2020 to May 20, 2021 | 277 | 638 | 45.8 | 29.8 | Not provided | 1 | 14 | RT-PCR | Community cases | Passive identification of secondary household cases independent from symptoms. | Likely all household unvaccinated |
| Metlay 2021^42^ | US | Retrospective cohort | Wild-type | March 4 - May 17, 2020 | 7,262 | 17,917 | Not stated | Not provided | 47.8 | Not stated | Not provided | RT-PCR | Community cases | Passive identification of secondary household cases independent from symptoms. | Likely all household unvaccinated |
| Miyahara, 2021^43^ | Japan | Cohort | Wild-type | February 22-May 31, 202 | 306 | 147 | 19 | Not provided | 49.7 | Not provided | Not provided | RT-PCR | Community cases | Passive identification of secondary cases independent from symptoms | Likely all households unvaccinated |
| Park 2020^26^ | South Korea | Surveillance data | Wild-type | January 20 to May 13 2020 | 5,706 | 59,073 | 11.8 | Not provided | Not provided | Multiple | 14 | RT-PCR | Community cases | Passive identification of secondary cases independent from symptoms | Likely all households unvaccinated |
| Rosenberg 2020^27^ | US | Surveillance data | Wild-type | March 2 – March 31, 2020 | 229 |  | Not stated | 43 | 43.7 |  |  | RT-PCR of throat swabs | Community cases | Passive identification of secondary cases independent from symptoms | Likely all households unvaccinated |
| Singanayagam 2021^18^ | UK | Prospective cohort | Wild-type, Alpha, Delta | September 13, 2020 to September 15, 2021 | 471 | 602 | 25 | 36 | 55 | Multiple | 14-20 | RT-PCR of throat swabs | Community cases | Active symptomatic follow-up, all contacts notified within 5 days of index case symptom | Fully vaccinated and unvaccinated individuals |
| Tibebu 2021^44^ | Canada | Surveillance data | Wild-type | July 1 to November 30, 2020 | 29,352 | 84,125 | 19.5 | 44 | 19 | 1 | 14-28 | RT-PCR | Community cases | Passive identification of secondary cases independent from symptoms | Likely all households unvaccinated |
| Wang, 2020^28^ | China | Retrospective case series | Wild-type | February 13-28 | 20,399 | 155 | 24 | Not provided | 48 | Multiple | Not stated | RT-PCR on throat swabs | Community cases | Passive identification of secondary cases independent from symptoms | Likely all households unvaccinated |
| Wei 2020^29^ | China | Surveillance data | Wild-type | Jan 1 to Feb 14, 2020 | 23 | 79 | 52 | 33.9 | 58.3 | Not stated | Not stated | Not provided | Community cases | Passive identification of secondary cases independent from symptoms | Likely all households unvaccinated |
| Wu, 2020^30^ | China | Surveillance data | Wild-type | January –February 2020 | 46 | 104 | 32.4 | 43.8 | Not provided | Not stated | 21 | RT-PCR of nasopharyngeal and/or oropharyngeal swabs | Community cases | Passive identification of secondary cases independent from symptoms | Likely all households unvaccinated |
| Yousaf 2020^45^ | US | Prospective cohort | Wild-type | 22 March to 22 April 2020 | 198 | 47 | Not stated | 24 | 62 | Multiple | 14 | RT-PCR of nasopharyngeal swab | Community cases | Active symptomatic follow-up | Likely all households unvaccinated |

**eTable 2: Risk of bias of included studies**

|  | **Representativeness of the index cases in region (2 points)^a^** | **Index case definition (1 point)^b^** | **Sample size (1 point)^c^** | **Household secondary attack rate disaggregated by index / contact age (1 point)^d^** | **Universal or symptomatic based testing (1 point)^e^** | **Follow-up duration ( 2 points)^f^** | **Number of test per contact (1 point)^g^** | **Total points** | **Risk of bias^h^** |
| --- | --- | --- | --- | --- | --- | --- | --- | --- | --- |
| Bi 2020^22^ | ++ | + | + | + | + | + | + | 8 | Low |
| Cerami 2021^31^ | + | + | 0 | + | + | ++ | + | 8 | Low |
| Ge 2021^32^ | ++ | + | + | + | + | ++ | + | 9 | Low |
| Grijalva 2020^16^ | + | + | + | + | + | ++ | + | 8 | Low |
| Jing 2020^23^ | ++ | + | + | + | + | ++ | + | 9 | Low |
| Julin 2021^68^ | + | + | 0 | + | + | ++ | + | 7 | Low |
| Koureas 2021^17^ | ++ | + | + | + | + | ++ | + | 9 | Low |
| Kuba 2021^34^ | + | + | 0 | + | + | ++ | 0 | 6 | Moderate |
| Lewis 2020^35^ | ++ | + | + | + | + | ++ | + | 9 | Low |
| Li 2020^24^ | + | + | 0 | + | + | ++ | + | 7 | Low |
| Li 2021^36^ | ++ | + | + | + | + | 0 | 0 | 6 | Moderate |
| Loenenbach 2021^37^ | + | + | 0 | + | + | ++ | 0 | 6 | Moderate |
| Loss, 2021^38^ | ++ | + | 0 | + | + | + | + | 7 | Moderate |
| Lyngse 2020^25^ | ++ | + | + | + | + | ++ | + | 9 | Low |
| Lyngse 2021^39^ | ++ | + | + | + | + | ++ | + | 9 | Low |
| Lyngse 2022^46^ | ++ | + | + | + | + | ++ | + | 9 | Low |
| Maltezoua 2021^40^ | + | + | + | + | + | ++ | 0 | 7 | Low |
| Martinez 2021^41^ | ++ | + | + | + | + | ++ | 0 | 8 | Low |
| Metlay 2021^42^ | ++ | + | + | + | + | 0 | 0 | 6 | Moderate |
| Miyahara, 2021^43^ | + | 0 | + | + | + | 0 | + | 5 | Moderate |
| Park 2020^26^ | ++ | + | + | + | + | ++ | + | 9 | Low |
| Rosenberg 2020^27^ | ++ | + | 0 | + | + | 0 | 0 | 5 | Moderate |
| Singanayagam 2021^18^ | + | + | + | + | + | ++ | + | 8 | Low |
| Tibebu 2021^44^ | ++ | + | + | + | + | ++ | 0 | 8 | Low |
| Wang, 2020^28^ | ++ | + | + | + | + | 0 | + | 7 | Low |
| Wei 2020^29^ | ++ | + | 0 | + | + | 0 | 0 | 5 | Moderate |
| Wu, 2020^30^ | + | + | 0 | + | + | ++ | 0 | 6 | moderate |
| Yousaf 2020^45^ | ++ | + | 0 | + | + | ++ | + | 8 | Low |

a ++: Representative of COVID-19 cases in region; +: Somewhat representative; 0: Poorly described or not representative of cases in region

b +: Index case identified by date of onset of symptoms and/or test dates; 0: First case not clearly defined

c +: ≥300 contacts; 0: <300 contacts

d +: Secondary attack rate disaggregated by ≥1 covariate; 0: Secondary attack rate not disaggregated by any covariates

e +: Tested all contacts (both symptomatic and asymptomatic); 0: Only tested symptomatic contacts

f ++: >14 days; +: 14 days; 0: <14 days or not specified

g +: ≥2 tests; 0: 1 test or not described

h High: ≤3 points; moderate: 4–6 points; low: ≥7 points

**Annex 1: Search strategy**

SARS-CoV-2/ or COVID-19/

(corona* adj1 (virus* or viral*)).mp.

(CoV not (Coefficien* or "co-efficien*" or covalent* or Covington* or covariant* or covarianc* or "cut-off value*" or "cutoff value*" or "cut-off volume*" or "cutoff volume*" or "combined optimi?ation value*" or "central vessel trunk*" or CoVR or CoVS)).mp.

(coronavirus* or 2019nCoV* or 19nCoV* or "2019 novel*" or Ncov* or "n-cov" or "SARS- CoV-2*" or "SARSCoV-2*" or SARSCoV2* or "SARS-CoV2*" or "severe acute respiratory syndrome*" or COVID*2).mp.

"Severe Acute Respiratory Syndrome Coronavirus 2".mp.

"COVID-19".mp.

"covid 19 diagnostic testing"2.mp.

"covid 19 drug treatment".mp.

"covid 19 serotherapy".mp.

"covid 19 vaccine".mp.

ncov*.mp.

covid*.mp.

sars-cov-2.mp.

"sars cov 2".mp.

"SARS Coronavirus 2".mp.

"Severe Acute Respiratory Syndrome CoV 2".mp.

"Wuhan coronavirus".mp.

"Wuhan seafood market pneumonia virus".mp.

"SARS2".mp.

"2019-nCoV".mp.

"hcov-19".mp.

"novel 2019 coronavirus".mp.

"2019 novel coronavirus*".mp.

"novel coronavirus 2019*".mp.

"2019 novel human coronavirus*".mp.

"human coronavirus 2019".mp.

"coronavirus disease-19".mp.

"corona virus disease-19".mp.

"coronavirus disease 2019".mp.

"corona virus disease 2019".mp.

"2019 coronavirus disease".mp.

"2019 corona virus disease".mp.

"novel coronavirus disease 2019".mp.

"novel coronavirus infection 2019".mp.

"new coronavirus*".mp.

"coronavirus outbreak".mp.

"coronavirus epidemic".mp.

"coronavirus pandemic".mp.

"pandemic of coronavirus".mp.

or/1-39

age.mp.

"age group*".mp.

Children*.mp.

child.mp.

childhood.mp.

teen*.mp.

pediatric*.mp.

paediatric*.mp.

adolescen*.mp.

boys.mp.

girls.mp.

youth.mp.

youths.mp.

or/41-53

(variant* or mutant* or mutation* or strain*).mp.

(alpha or S-gene target failure or SGTF or beta or gamma or delta or epsilon or zeta or eta or theta or iota or kappa or lambda).mp.

("B.1.1.7" or "20I/501Y.V1" or "VOC 202012/01" or "B.1.351" or "20H/501Y.V2" or "P.1" or "20J/501Y.V3" or "B.1.1.28" or "501Y*" or "N501Y" or "E484K" or "D614G" or "69/70 deletion" or "144Y deletion" or "A570D" or "P681H" or "K417N*" or "VUI-202012/01" or "Kent" or "B.1.427" or "CA VUI1" or "CAL.20C" or "B.1.429" or "B.1.526" or "B.1.525" or "P.2" or "P.3" or "B.1.617" or "B.1.617.2" or "C.37").mp.

or/55-57

transmission.mp.

disease susceptibility.mp.

communicability.mp.

contagious.mp.

contagiousness.mp.

susceptibility.mp.

epidemiology.mp.

"contact tracing".mp.

"communicable disease contact tracing".mp.

infection.mp.

infectious*.mp.

"attack rate".mp.

“household”.mp.

"secondary attack rate".mp.

or/59-72

(shed* or viabl*).mp.

(viral or virus or rna or ribonucleic).mp.

viral clearance.mp.

viral load.mp.

viral shedding.mp.

or/74-78
